# Ordinal regression increases statistical power to predict epilepsy surgical outcomes

**DOI:** 10.1101/2021.10.01.21264435

**Authors:** Adam S. Dickey, Robert T. Krafty, Nigel P. Pedersen

## Abstract

**Objective:** Studies of epilepsy surgery outcomes are often small and thus underpowered to reach statistically valid conclusions. We hypothesized that ordinal logistic regression would have greater statistical power than binary logistic regression when analyzing epilepsy surgery outcomes.

**Methods:** We reviewed 10 manuscripts included in a recent meta-analysis which found that mesial temporal sclerosis (MTS) predicted better surgical outcome after a stereotactic laser amygdalohippocampectomy (SLAH). We extracted data from 239 patients from eight studies which reported four discrete Engel surgical outcomes after SLAH, stratified by the presence or absence of MTS.

**Results:** The rate of freedom from disabling seizures (Engel I) was 64.3% (110/171) for patients with MTS compared to 44.1% (30/68) without MTS. The statistical power to detect MTS as a predictor for better surgical outcome after a SLAH was 29% using ordinal regression, which was significantly more than the 13% power using binary logistic regression (paired t-test, p<0.001). Only 120 patients are needed to achieve 80% power to detect MTS as a predictor using ordinal regression, compared to 210 patients that are needed to achieve 80% power using binary logistic regression.

**Conclusion:** Ordinal regression should be considered when analyzing ordinal outcomes (such as Engel surgical outcome), especially for datasets with small sample sizes.

## INTRODUCTION

Studies of epilepsy surgery often aim to identify predictors of good outcome to help guide treatment^1^ However, most studies treat seizure outcome as a binary variable of seizure freedom (Engel I) or not, which ignores the difference between rare disabling seizures (Engel II), worthwhile improvement in seizure frequency (Engel III), and no worthwhile improvement (Engel IV)^2^. We argue that statistical analysis should leverage the full range of possible outcomes.

We propose using *ordinal logistic regression* rather than traditional binary logistic regression, as the Engel surgical outcome is an ordinal scale. Relying on seizure freedom as a primary outcome amounts to comparing Engel I vs. Engel II-IV. There are two additional dichotomous categorizations: Engel I-II vs. Engel III-VI and Engel I-III vs. Engel IV. Rather than fit three individual binary regressions, ordinal regression fits three separate intercepts and a common, pooled odds ratio^3^. The additional assumption required is that the odds ratio is similar between the three comparisons.

It has been shown that binarizing an ordinal categorical outcome can lead to a reduction in statistical power in some settings^4, 5^. We hypothesis than ordinal logistic regression using full Engel outcomes, by discarding less outcome information, will have more statistical power than binary logistic regression. We test this hypothesis using the problem of predicting surgical outcome after stereotactic laser amygdalohippocampectomy (SLAH). A recent meta-analysis of 10 studies found that the presence of mesial temporal sclerosis (MTS) predicted a better chance (63% vs. 42%) of seizure freedom after SLAH when MTS was present than when absent^6^. However, most individual studies did not detect this effect, as their low sample size limited statistical power. We specifically test whether ordinal logistic regression has more statistical power than binary logistic regression to detect the known relation between MTS and seizure outcome in each individual study.

## METHODS

A recent meta-analysis included 10 studies in its comparison of seizure freedom in the presence or absence of MTS^6^. We reviewed the original manuscripts and found eight studies that reported raw counts of patients for each of the four discrete Engel outcomes, stratified by presence or absence of MTS^7–14^. We excluded one paper because it used an alternative seizure outcome scale^15^ and another because it grouped together the classification of Engel II-IV^16^. This study is thus a meta-analysis of publicly available, published studies that were approved by their local Institutional Review Board (IRB) or its equivalent.

### Logistic regression

For each study, we converted the count data into an independent vector with the presence (1) or absence (0) of MTS, and dependent vectors of seizure freedom (1) or not (2), or the full Engel outcomes (1-4). We performed binary logistic regression and ordinal logistic regression on the dependent vectors of seizure freedom or Engel outcome, respectively. We report the p-value for the log-odds (or slope) coefficient. Both the binary and ordinal regression were performed using the Matlab function **mnrfit** in MATLAB (version R2016b, Mathworks, Natick, MA) using a parallel ordinal model, which assumes a common odds ratio. When this function is supplied a vector with two outcomes, it provides the same results as binary logistic regression.

### Power analysis

We pooled the count data across the eight studies to compute an empiric distribution of the probability of each Engel outcome in the presence or absence of MTS. We performed a power analysis by simulating binomial count data, using the empiric probability distribution and the count data of patients with or without MTS from each individual study. We performed a bootstrap simulation with 10,000 repetitions and report the power as the percentage of repetitions where the logistic regression (binary or ordinal) reported a p-value for the log-odds coefficient as less than 0.05.

## RESULTS

The raw and pooled count data are summarized below (Table 1). Across the eight studies, 72% (171 /239) of patients had MTS and 28% (68/239) did not. The rate of freedom from disabling seizures (Engel I) was 64.3% (110/171) for patients with MTS compared to 44.1% (30/68) without MTS. These results are similar to those from the meta-analysis (63% vs. 42%), which included 10 studies^6^.

**Table 1:**
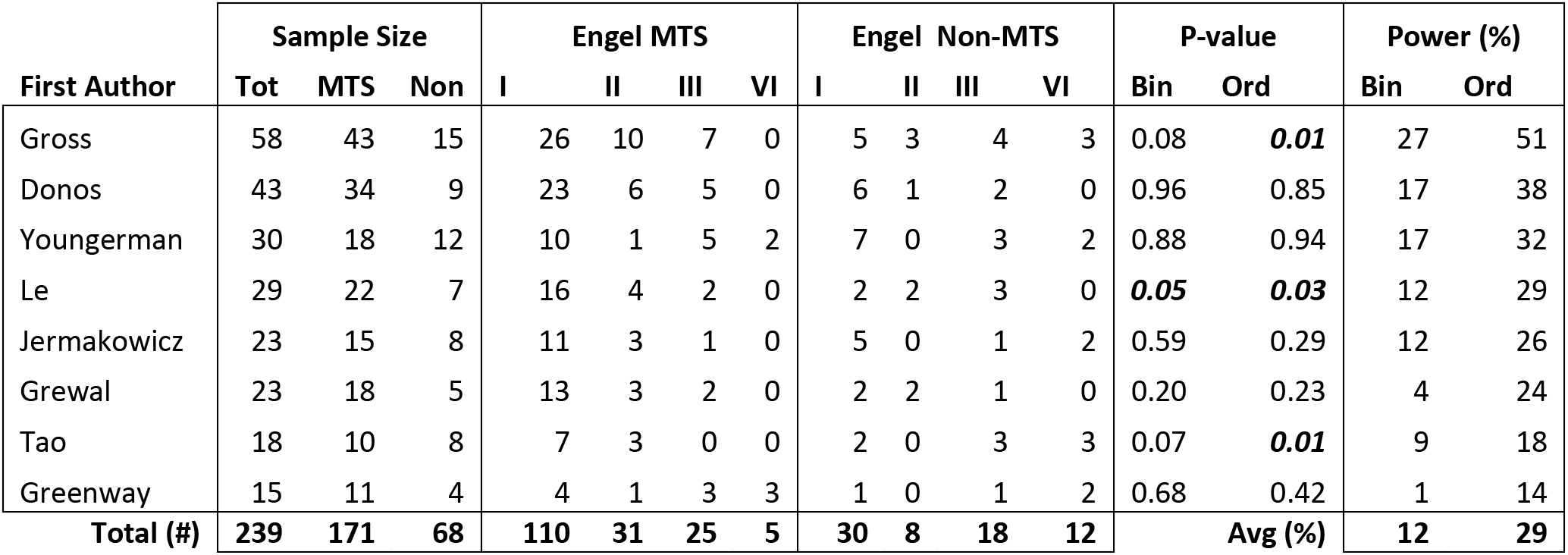
Patient counts by study for Engel outcome for mesial temporal sclerosis or not. Total = total; MTS = mesial temporal sclerosis; Non = non-MTS; Engel = Engel surgical outcome; Bin = binary logistic regression; Ord = Ordinal logistic regression. Avg = Average. P-values < 0.05 are italicized and bolded.

A comparison of the observed distribution of Engel outcomes to that predicted by ordinal regression is shown, stratified by the presence or absence of MTS (Figure 1). The pooled odds ratio assumption means the predicted difference in Engel I for MTS or not (65% vs. 39%) is slightly higher than observed (64 vs. 44%), but the predicted difference for Engel IV for MTS or not (5% vs. 13%) is smaller than observed (3% vs. 18%). A comparison of the pooled odds ratio and the odds ratios of the three possible binary logistic regressions is also shown (Figure 2). The pooled odds ratio of 2.97 is contained within the 95% confidence interval for each binary logistic regression, so the pooled odds ratio assumption does not appear to be violated.

**Figure 1:**
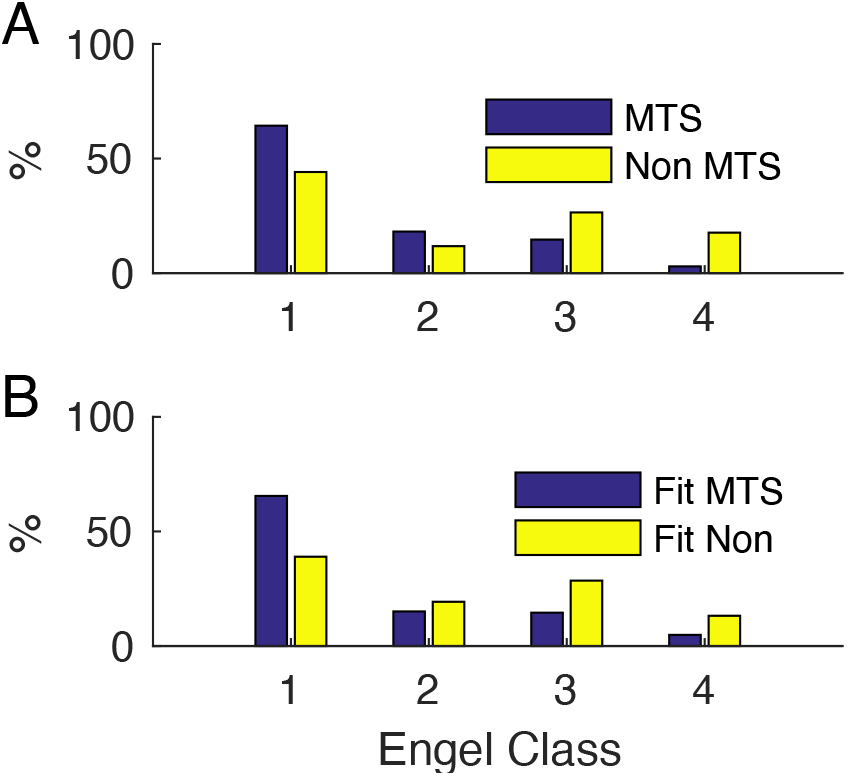
Results of ordinal logistic regression. (A) Observed percentages of Engel outcomes I-IV stratified by presence or absence of mesial temporal sclerosis (MTS), pooled from 8 studies. (B) Predicted percentages of Engel I-VI stratified by MTS or not, using ordinal logistic regression. The pooled odds ratio assumption means the predicted difference in Engel I for MTS or not (65% vs. 39%) is slightly higher than observed (64 vs. 44%), but the predicted difference for Engel IV for MTS or not (5% vs. 13%) is smaller than observed (3% vs. 18%).

**Figure 2:**
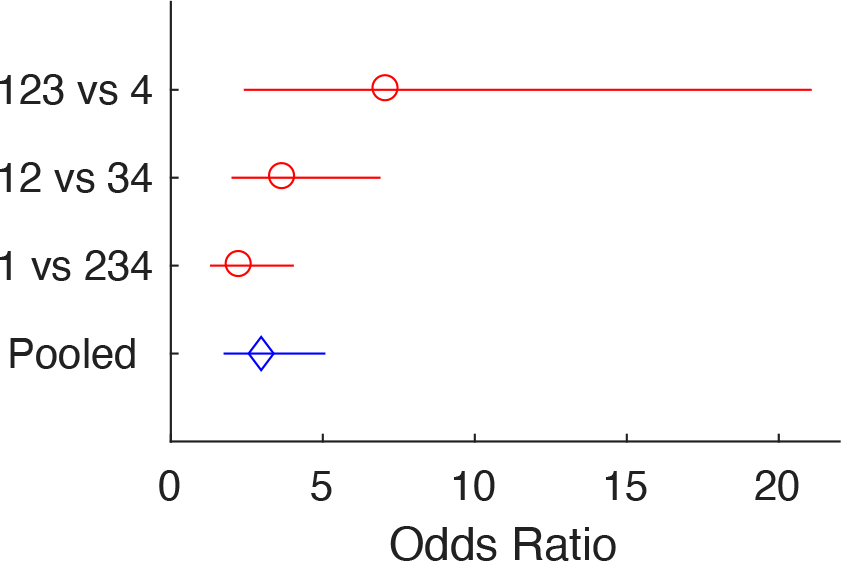
Odds ratios for binary vs. pooled ordinal logistic regression. Predicted odds ratios (with 95% confidence interval) are display for the three possible binary logistic regressions, compared to the pooled odds ratio from ordinal logistic regression. The pooled odds ratio of 2.97 is contained within the 95% confidence interval for each binary logistic regression, so the pooled odds ratio assumption does not appear to be violated.

### Statistical significance

When the raw count data was analyzed for each study using binary logistic regression, only one study^12^ showed a significant difference (p < 0.05) for seizure freedom with versus without MTS. When ordinal logistic regression was used, three studies showed a significant difference in Engel outcome for patients with versus without MTS. Of note, both studies with a significant result for ordinal, but not binary regression, did show a significant difference related to MTS using a Kaplan-Meier analysis. However, those analyses included extended follow up to 53 months^10^ and 43 months^13^.

### Power Analysis

We used the pooled count data to estimate an empirical distribution of the probability of each Engel outcome given presence or absence of MTS (Table 1). We assumed this as ground truth and generated simulated data for each study, given the number of subjects with or without MTS. We then estimated the power to detect a difference in Engel outcome using binary or ordinal logistic regression. The mean statistical power to detect the true effect was 29% for ordinal regression, which was significantly more than 13% for binary logistic regression (paired t-test, p<0.001).

We estimated the sample size needed to achieve 80% power to detect a difference in Engel outcome, assuming a 2:1 allocation of groups (two-thirds with MTS and one third without). Using binary logistic regression (or a chi-square test), one would need around 210 patients (140 with MTS, 70 without) to achieve 80% power to detect a difference of seizure freedom of 64.3% vs. 44.1% (boot-strap, 10,000 repetitions).

However, using a pooled empiric probability distribution for Engel outcomes (Table 1), one achieves 80% power to detect the same difference in Engel outcome with only 120 patients (boot-strap, 10,000 repetitions, 80 with MTS, 40 without). In contrast, binary logistic regression and the chi-squared test only have ∼56% power with 120 patients. For this example, switching from binary to logistic regression almost doubles the effective sample size.

## DISCUSSION

To our knowledge, this is the first published report applying ordinal logistic regression Engel surgical outcomes. Ordinal regression increases statistical power and decreases the sample size need to achieve a desired power. Though we use data from multiple studies, one limitation is that we focus on a specific problem in epilepsy surgery, so the generalizability of this finding is not yet clear.

Because SLAH is a relatively new technique, cases series necessarily have small sample sizes and thus low statistical power. Underpowered studies will miss true effects when present (such as the individual studies which did not find an association between MTS and seizure outcome). Underpowered studies also have inflated false positive rates^17^ and overestimate the magnitude of statistically significant effects when found^18^. Low powered studies distort our understanding of prognostics factors for epilepsy surgery.

The obvious solution is to acquire larger sample sizes. However, this is difficult in practice. The less obvious solution is to use more sophisticated statistical analysis to avoid discarding relevant information^3^. Here, binary classification of seizure freedom is suboptimal when full Engel outcomes are available. It may also be possible to increase statistical power by using continuous predictors. For example, asymmetry scores for MTS could be computed from neuroimaging data, similar to those used for WADA tests^19^.

Our motivation for this paper was not re-demonstrating that MTS predicts seizure freedom after SLAH. Rather, the goal is identifying statistical tools which can detect relevant prognostic factors in small datasets. For example, semiology has been under-analyzed as a predictive factor for good outcome after SLAH. Signs such as ipsilateral manual automatisms and contralateral dystonia are strongly lateralizing^20^. Well-selected patients with concordant semiology (and other presurgical data) may have better surgical outcomes that patients with MTS but discordant semiology or scalp EEG. Ordinal regression should be an ideal tool to clarify the relative predictive value of semiology versus neuroimaging findings.

Ordinal regression is applicable outside of epilepsy as well. One paper showed that ordinal regression increased the power to detect the beneficial effect of corticosteroids on the Glasgow outcome scale in patients with traumatic brain injury^5^. We expect it would be applicable to the ordinal modified Rankin scale, which is often used in stroke literature. In summary, ordinal regression should always be considered when analyzing ordinal outcomes, especially for datasets with small sample sizes.

## Data Availability

MATLAB code which can be used to reproduce the analyses and figures described here are posted at https://github.com/AdamSDickey/Ordinal_Regression.

## FUNDING AND ACKNOWLEDGEMENTS

N.P.P. is supported by the Woodruff Foundation, CURE Epilepsy, and NIH grants K08 NS105929, R01 NS088748, and R21 NS122011. A.S.D. is supported by the National Center for Advancing Translational Sciences of the NIH under award number UL1 TR002378 and KL2 TR002381. R.T.K. is supported by R01 GM113243. The content is solely the responsibility of the authors and does not necessarily represent the official views of the National Institutes of Health. We also thank Scott Millis of Wayne State University for help finding relevant references.

## AUTHORSHIP STATEMENTS

A.S.D and N.P.P. contributed to the conception and design of the study. A.S.D, R.T.K and N.P.P contributed to the drafting of the text. A.S.D. performed the statistical analysis and prepared the figures and tables

## REFERENCES

1. Jobst BC, Cascino GD. Resective epilepsy surgery for drug-resistant focal epilepsy: a review. JAMA. 2015 Jan 20;313(3):285–93.

2. Engel J. Update on surgical treatment of the epilepsies: summary of the second international palm desert conference on the surgical treatment of the epilepsies (1992). Neurology. 1993;43(8):1612–.

3. Steyerberg EW. Clinical prediction models: Springer; 2019.

4. Whitehead J. Sample size calculations for ordered categorical data. Statistics in medicine. 1993;12(24):2257–71.

5. Roozenbeek B, Lingsma HF, Perel P, et al. The added value of ordinal analysis in clinical trials: an example in traumatic brain injury. Critical Care. 2011;15(3):1–7.

6. Kohlhase K, Zollner JP, Tandon N, Strzelczyk A, Rosenow F. Comparison of minimally invasive and traditional surgical approaches for refractory mesial temporal lobe epilepsy: A systematic review and meta-analysis of outcomes. Epilepsia. 2021 Apr;62(4):831–45.

7. Donos C, Breier J, Friedman E, et al. Laser ablation for mesial temporal lobe epilepsy: Surgical and cognitive outcomes with and without mesial temporal sclerosis. Epilepsia. 2018 Jul;59(7):1421–32.

8. Greenway MRF, Lucas JA, Feyissa AM, Grewal S, Wharen RE, Tatum WO. Neuropsychological outcomes following stereotactic laser amygdalohippocampectomy. Epilepsy Behav. 2017 Oct;75:50–5.

9. Grewal SS, Zimmerman RS, Worrell G, et al. Laser ablation for mesial temporal epilepsy: a multisite, single institutional series. J Neurosurg. 2018 Jul 1:1–8.

10. Gross RE, Stern MA, Willie JT, et al. Stereotactic laser amygdalohippocampotomy for mesial temporal lobe epilepsy. Ann Neurol. 2018 Mar;83(3):575–87.

11. Jermakowicz WJ, Kanner AM, Sur S, et al. Laser thermal ablation for mesiotemporal epilepsy: Analysis of ablation volumes and trajectories. Epilepsia. 2017 May;58(5):801–10.

12. Le S, Ho AL, Fisher RS, et al. Laser interstitial thermal therapy (LITT): Seizure outcomes for refractory mesial temporal lobe epilepsy. Epilepsy Behav. 2018 Dec;89:37–41.

13. Tao JX, Wu S, Lacy M, et al. Stereotactic EEG-guided laser interstitial thermal therapy for mesial temporal lobe epilepsy. J Neurol Neurosurg Psychiatry. 2018 May;89(5):542–8.

14. Youngerman BE, Oh JY, Anbarasan D, et al. Laser ablation is effective for temporal lobe epilepsy with and without mesial temporal sclerosis if hippocampal seizure onsets are localized by stereoelectroencephalography. Epilepsia. 2018 Mar;59(3):595–606.

15. Kang JY, Wu C, Tracy J, et al. Laser interstitial thermal therapy for medically intractable mesial temporal lobe epilepsy. Epilepsia. 2016 Feb;57(2):325–34.

16. Waseem H, Vivas AC, Vale FL. MRI-guided laser interstitial thermal therapy for treatment of medically refractory non-lesional mesial temporal lobe epilepsy: Outcomes, complications, and current limitations: A review. J Clin Neurosci. 2017 Apr;38:1–7.

17. Button KS, Ioannidis JP, Mokrysz C, et al. Power failure: why small sample size undermines the reliability of neuroscience. Nat Rev Neurosci. 2013 May;14(5):365–76.

18. Dickey AS, Pedersen NP. Low statistical power in a study predicting seizure outcome. Epilepsia. 2021 Aug 5.

19. Loring DW, Bowden SC, Lee GP, Meador KJ. Diagnostic utility of Wada Memory Asymmetries: sensitivity, specificity, and likelihood ratio characterization. Neuropsychology. 2009 Nov;23(6):687–93.

20. Kotagal P, Lüders H, Morris H, et al. Dystonic posturing in complex partial seizures of temporal lobe onset: a new lateralizing sign. Neurology. 1989;39(2):196–.

